# Characterization of metabolically healthy and unhealthy obesity through circulating proteins and metabolites

**DOI:** 10.1101/2025.03.10.25323679

**Authors:** Giulia Pontali, Christian X. Weichenberger, Johannes Rainer, Essi Hantikainen, Marilyn De Graeve, Fulvio Mattivi, Michael Kob, Markus Ralser, Peter P. Pramstaller, Francisco S. Domingues

## Abstract

**Background:** Individuals affected by obesity present different health trajectories and do not suffer from cardiometabolic complications all in the same way. There is a need to better understand obesity subtypes and to develop approaches for stratification. In this study we investigated both metabolomic and proteomic signatures in serum and blood plasma samples discriminating metabolically healthy from unhealthy obesity.

**Methods:** We investigated cross-sectional metabolomic and proteomic data from participants of the Cooperative Health Research in South Tyrol (CHRIS) study. Participants were grouped into metabolically healthy obesity (MHO) and metabolically unhealthy obesity (MUO) based on available health data in the study. A total of 461 individuals were included in the analysis, with n=130 MHO and n=331 MUO. Random forest (RF) classifiers were used to discriminate metabolically healthy from unhealthy obesity and to identify molecular features characteristic of MHO/MUO. Linear regression models were used to assess associations between each relevant metabolite/protein and MHO/MUO phenotypes independently of age, sex and body composition.

**Results:** The MHO/MUO RF classifier achieved a performance of AUC = 0.709, 95% CI = (0.698,0.721). Three plasma proteins and 12 circulating metabolites were identified as relevant predictors of MHO/MUO phenotypes. Linear regression models confirmed the Apolipoprotein C-III (APOC3) association to be independent of age, visceral fat composition, medication or serum triglyceride levels.

**Conclusion:** APOC3 was identified as a novel predictor for obesity stratification, highlighting the importance of circulating triglyceride levels in relation to metabolic health.

## INTRODUCTION

Obesity is a condition characterized by abnormal accumulation of body fat, that has been linked to type 2 diabetes, cardiovascular disease, and some types of cancer [1, 2]. It has been estimated that overweight and obesity accounts for a considerable proportion of chronic disease burden and premature death [3–5]. The global prevalence of obesity has been increasing. Recent estimates indicate that between 1990 and 2022 obesity prevalence in adults increased from 8.8% to 18.5% in women, and from 4.8% to 14.0% in men, overtaking the global prevalence of underweight in both genders in this period [6].

Obesity results from an interplay between genetics, lifestyle (specifically diet) and environmental factors [7], and is widely regarded as a multifactorial disease [8]. There is a substantial genetic component underlying individual variation in body weight, and different studies have estimated the heritability of obesity to be between 40%-70% [9]. Genetic studies also indicate that monogenic and polygenic obesity share a similar biology, where central nervous system pathways related to food intake play a key role [9]. Furthermore, the gut microbiome impacts obesity by modulating mood disorders, eating behaviors and detoxification processes [10].

Not all individuals affected by obesity suffer from metabolic complications, which cannot be explained only by the extent of adiposity. A considerable subgroup of individuals with obesity seems to exhibit a relative protection or a significantly lower risk of cardiometabolic disease. This phenotype has been described as metabolically healthy obesity (MHO), to differentiate from the more prevalent metabolically unhealthy obesity (MUO) [11]. Multiple definitions for MHO/MUO have been proposed, but in general they include the body mass index (BMI) criteria for obesity, and the absence/presence of hyperglycemia, dyslipidemia, hypertension and cardiovascular disease [11, 12]. MUO has been linked to higher visceral body fat, a proinflammatory state and insulin resistance, while MHO is characterized by a higher subcutaneous body fat, greater insulin sensitivity and better cardiorespiratory fitness [11, 13]. MUO is potentially the outcome of a long-term positive energy balance coupled with inability of subcutaneous adipose tissue to expand [14–16]. Although MHO is associated with a lower risk of cardiometabolic disease in comparison to MUO, it has long-term negative impact on cardiometabolic health and should therefore not be considered a benign condition [17, 18]. To some extent MHO is a transient phenotype to MUO, although the MHO phenotype can be stable over years in a subset of individuals [19–22]. However, the transition to MUO is not only unidirectional, as conversion from MUO to MHO is also being observed [23].

High-throughput omics approaches capture detailed molecular profiles in biological samples and allow us to investigate any relevant changes between health conditions, providing insights into the relevant processes underlying health and disease [24]. With such detailed molecular profiling is then possible to better characterize disease subtypes, enabling enhanced stratification of individuals for improved diagnosis, prognosis and more effective therapeutic strategies [25]. These approaches have been successfully applied in the investigation of metabolic health in obesity [26]. Extensive metabolic profiling of the MHO phenotype [27–34] has highlighted differences between MHO and MUO in the levels of several metabolites and identified altered metabolic pathways. The results indicate differences in the levels of branched-chain amino acids and aromatic amino acids, where the blood amino acid profile of MHO falls between those of metabolically healthy individuals with normal weight and MUO [26].

Circulating proteomics profiles for MHO/MUO have also been investigated, with results pointing towards increased inflammation and insulin resistance in MUO [35–38]. These proteomics studies are based on relatively modest sample sizes, limiting the power to determine relevant features for MHO and for stratification of obesity.

In this study, we perform a large-scale analysis of serum metabolomic and plasma proteomic signatures in MHO and MUO, utilizing the largest sample size for combined omics analysis reported to date. This study aims to identify key molecular features that differentiate these conditions, contributing to more precise stratification of obesity.

## METHODS

### Study Cohort

The Cooperative Health Research in South Tyrol (CHRIS) study is a population-based cohort of 13,393 adults aged 18 and over, recruited from 13 municipalities in the alpine Val Venosta/Vinschgau district in the Bolzano-South Tyrol province of northern Italy [39]. Baseline visits were conducted from 2011 to 2018, collecting socio-demographic, health, lifestyle, medication and exposure data with questionnaires, interviews, and instrumental examinations, including electrocardiography, blood pressure and anthropometric measurements. Body composition, including body fat and visceral fat percentages were assessed by bioelectrical impedance (OMRON BF508). Medication information was collected by scanning the barcode of the boxes of the medication taken over the last seven days by each study participant and used to assign the respective Anatomical Therapeutic Chemical (ATC) codes. Blood and urine samples were also collected for biobanking, DNA extraction, and molecular characterization (genome and exome sequencing, metabolomics, and proteomics). The CHRIS resource was assigned the “Bioresource Research Impact Factor” (BRIF) code BRIF6107. Standard blood parameters were measured in blood samples at the Hospital of Merano using standardized clinical assays as described previously [40]. More specifically, the serum triglycerides were measured either with Cobas Triglyceride GPO-PAP assay on a Roche Modular PPE instrument or with a TRIGLYCERIDE assay on an Abbot Diagnostic ARCHITECT system.

### Assessment of Obesity Phenotypes

Obesity was defined as having a BMI greater than or equal to 30 kg/m^2^ [41]. Metabolically healthy obesity (MHO) and metabolically unhealthy obesity (MUO) were defined according to previously established criteria [42] as summarized in Table 1. Briefly, MUO is defined as individuals with obesity that are affected by either hypertension, diabetes, dyslipidemia or cardiovascular disease, while MHO is defined as all other individuals with obesity that are not MUO and where metabolic health could be assessed (no missing data). The two phenotypes were encoded in a single binary variable (MHO/MUO phenotypes).

**Table 1.**
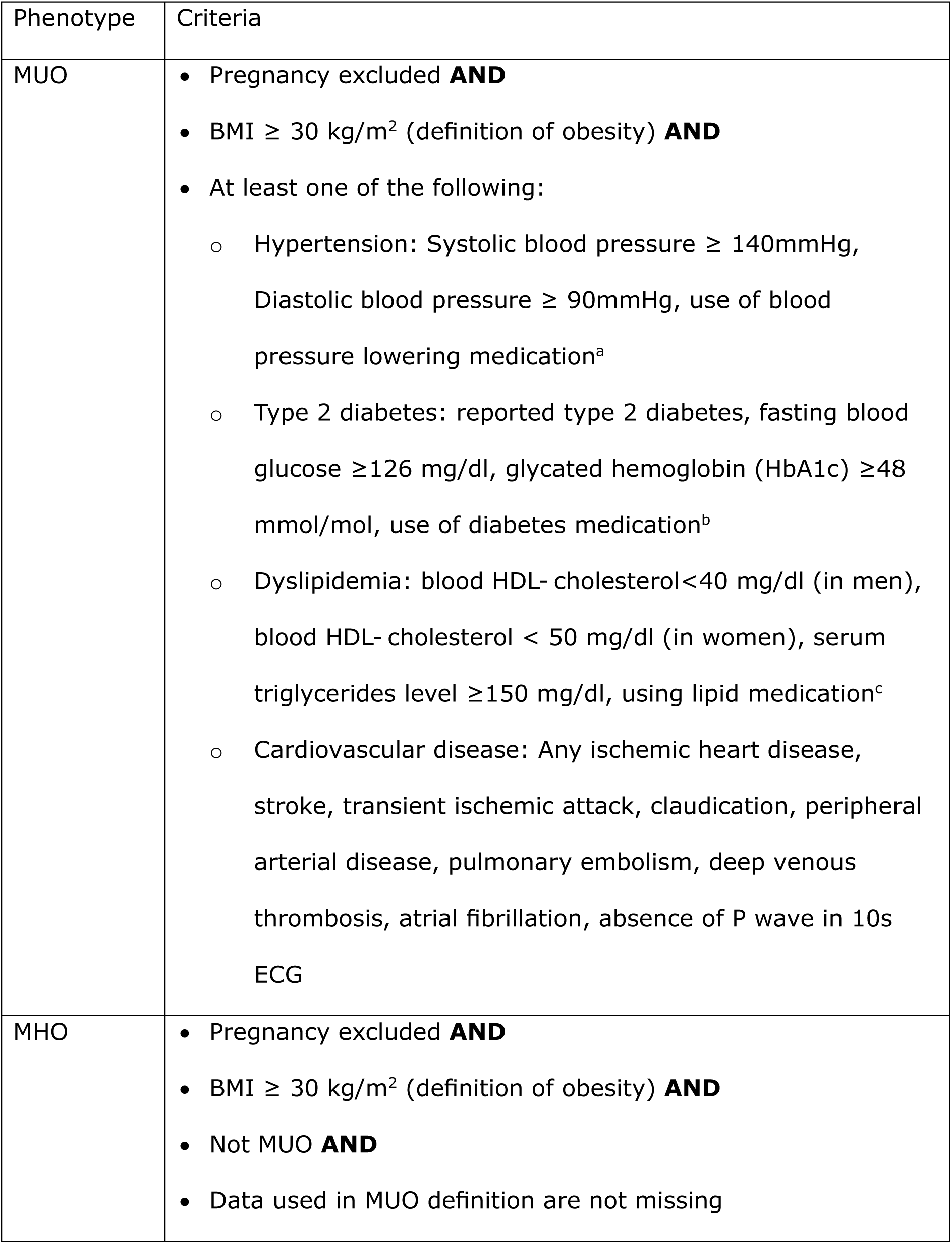
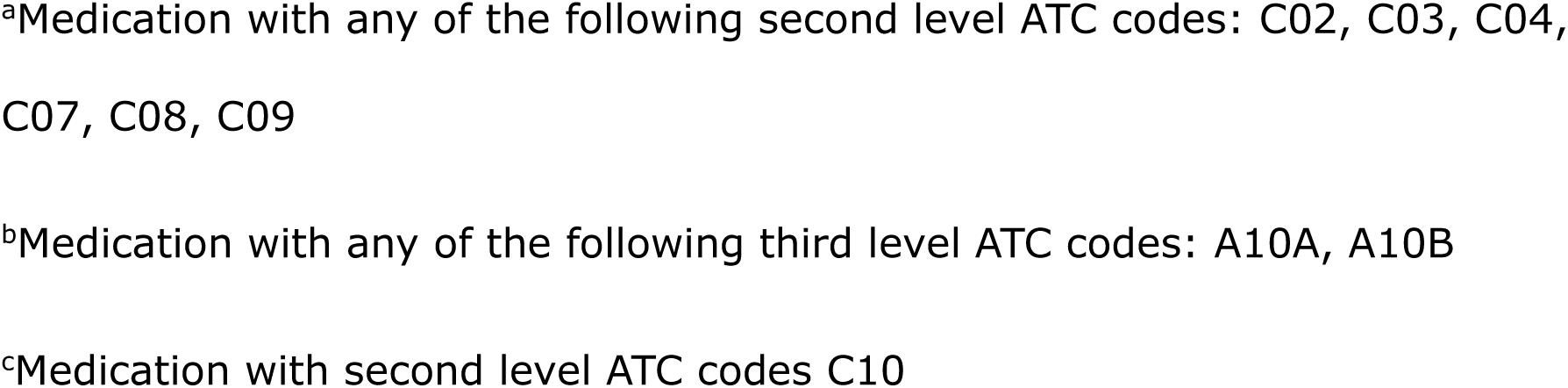
Criteria used in definition of MHO and MUO.

| Phenotype | Criteria |
| --- | --- |
| MUO | • Pregnancy excluded **AND**<br>• BMI $\geq 30 \text{ kg/m}^2$ (definition of obesity) **AND**<br>• At least one of the following:<br>○ Hypertension: Systolic blood pressure $\geq 140\text{mmHg}$, Diastolic blood pressure $\geq 90\text{mmHg}$, use of blood pressure lowering medication<sup>a</sup><br>○ Type 2 diabetes: reported type 2 diabetes, fasting blood glucose $\geq 126 \text{ mg/dl}$, glycated hemoglobin (HbA1c) $\geq 48 \text{ mmol/mol}$, use of diabetes medication<sup>b</sup><br>○ Dyslipidemia: blood HDL- cholesterol $< 40 \text{ mg/dl}$ (in men), blood HDL- cholesterol $< 50 \text{ mg/dl}$ (in women), serum triglycerides level $\geq 150 \text{ mg/dl}$, using lipid medication<sup>c</sup><br>○ Cardiovascular disease: Any ischemic heart disease, stroke, transient ischemic attack, claudication, peripheral arterial disease, pulmonary embolism, deep venous thrombosis, atrial fibrillation, absence of P wave in 10s ECG |
| MHO | • Pregnancy excluded **AND**<br>• BMI $\geq 30 \text{ kg/m}^2$ (definition of obesity) **AND**<br>• Not MUO **AND**<br>• Data used in MUO definition are not missing |

<sup>a</sup>Medication with any of the following second level ATC codes: C02, C03, C04, C07, C08, C09
<sup>b</sup>Medication with any of the following third level ATC codes: A10A, A10B
<sup>c</sup>Medication with second level ATC codes C10

### Metabolite and protein quantification

Metabolomics and proteomics data were collected as previously described [43]. To summarize, the AbsoluteIDQ® p180 kit from Biocrates (Biocrates Life Sciences AG, Innsbruck, Austria) was used for metabolite quantification in serum samples collected in fasting participants. Samples were analyzed with a UHPLC-MS/MS system consisting of an Agilent 1290 Infinity liquid chromatography system (Agilent Technologies, CA, USA) and a Sciex QTRAP 6500 mass spectrometer (AB Sciex LLC, MA, USA), and data were processed as described previously [44]. In total, 174 metabolites were quantified, they are provided in Supplementary Table 1. The plasma proteome was determined using Scanning SWATH mass spectrometry [45] as described previously [46]. In total, 148 highly abundant proteins were quantified and included in this analysis, they are listed in Supplementary Table 2.

### Medication Adjustment

Metabolite and protein abundances were log_2_-transformed and adjusted for frequent medication as previously described [43]. More specifically, linear models were fitted separately for each metabolite or protein with their abundance as response, and medication (encoded as individual binary variables) as explanatory variables. Only medication taken at least twice per week was considered for the adjustment. Medication used for MHO/MUO definitions were excluded (blood pressure, diabetes or blood lipid medication). The residuals from these models were used to construct medication-independent metabolite and protein abundances for all subsequent analyses. Supplementary Table 3 lists the types of medication for which abundances were adjusted, and the linear models are described in Supplementary file section 4.1.

### Random forest analysis

A random forest (RF) classification model was used to predict MHO vs. MUO, encoded as a binary variable (MHO/MUO phenotypes), and following a similar strategy to a previous analysis [43] using as predictors age, sex, body fat, visceral fat, 174 metabolites and 148 proteins. Overall, 100 RF models were generated, each containing 500 trees per model, with 80% training set sizes. Predictions were validated 100 times by repeated random subsampling with 20% test set sizes, while keeping the same ratio of MHO to MUO cases in the original and subsampled sets. The statistical programming language R v4.2.0 was utilized for computations and figure creation. More specifically, the randomForest R package v4.7-1.1 [47] was used to perform the RF analysis. Model performance was measured based on the receiver operating characteristic (ROC) area under the curve (AUC), and the Matthew’s correlation coefficient (MCC). The MCC was calculated at the optimal threshold on the ROC curve, corresponding to the true positive rate/false positive rate pair most distant from the diagonal. The AUC and MCC values reported correspond to the means of the 100 test runs, provided with the respective 95% confidence interval (CI). The Gini Index was used to measure feature importance. The significance of the Gini Indices for the random forest models was estimated by permuting the response variable using the rfPermute R package v2.5.1 [48]. This generates a background distribution by calculating 100 random forests, one for each permuted response variable, each with 500 trees. A background distribution for each validation run was created, yielding 100 p-values for each predictor variable. If in ≥ 50 out of the 100 models the predictor was significant at the level α = 0.05 it was identified as a relevant predictor.

### Confirmation of relevant predictors

Linear regression models were applied to further investigate associations between MHO/MUO and the relevant metabolites or proteins obtained from the RF analysis. Separate multiple linear regression models were fitted with abundances of each metabolite and protein as the response variable, and with MHO/MUO status, age, sex and visceral fat as the explanatory variables (Supplementary file section 4.2). The resulting p-values were adjusted for multiple hypothesis testing using the Bonferroni method. After correction, results were considered statistically significant if the adjusted p-value remained below the conventional significance level of α = 0.05. Additional linear models were derived to investigate associations between relevant proteins and metabolites and medication used in phenotype definition (Supplementary file section 4.3). Logistic regression models were used to investigate if the association between MHO/MUO phenotypes and plasma APOC3 is dependent on serum triglyceride levels (Supplementary file section 4.4).

### Ethical approval

The Ethics Committee of the Healthcare System of the Autonomous Province of Bolzano-South Tyrol approved the CHRIS baseline protocol on 19 April 2011 (21-2011). The CHRIS study conforms to the Declaration of Helsinki, and with national and institutional legal and ethical requirements.

## RESULTS

In this study, serum metabolomic and plasma proteomic signatures discriminating MHO from MUO are investigated on 461 subjects of the CHRIS study. First, the main characteristics of the study samples are provided. Second, the results from the random forest (RF) analysis are provided, including the prediction performance and the identification of relevant predictors for the classification of MHO/MUO. Third, the results of the multiple linear regression analyses are presented to support the identification of molecular predictors discriminating MHO from MUO.

### Characteristics of the study sample

Data were available in the CHRIS study to assign obesity status for a total of 13,216 participants, with 2,238 of them being affected by obesity (16.93%). The data required to assign MHO/MUO phenotypes were available for a subset of this obesity group (n=2,170), and among these a total of 529 participants were classified as MHO (24.38%) and 1,641 as MUO (75.62%). All the subsequent analyses are based on subsets with available metabolomics and proteomics data, for a total size of n=130 for MHO and n=331 MUO groups. The characteristics of this analytic sample are provided in Table 2. A lower mean age and a larger proportion of females are observed in the MHO group in comparison to MUO, and hypertension is the most frequent condition in the MUO group.

**Table 2.** Characteristics of the analytic sample grouped by metabolically healthy (MHO) and unhealthy obesity (MUO) *.

| Characteristic | MHO (n=130) | MUO (n=331) |
| --- | --- | --- |
| Age (years) | 43.98 ± 13.94 | 57.10 ± 14.73 |
| Male | 47 (36.15%) | 165 (49.85%) |
| Female | 83 (63.85%) | 166 (50.15%) |
| BMI (kg/m<sup>2</sup>) | 32.98 ± 3.14 | 33.43 ± 3.10 |
| Visceral fat (%) | 12.09 ± 4.07 | 15.21 ± 4.08 |
| Body fat (%) | 41.19 ± 8.62 | 39.65 ± 8.34 |
| Hypertension | 0 (0.00%) | 245 (74.02%) |
| Type 2 diabetes | 0 (0.00%) | 58 (17.52%) |
| Dyslipidemia | 0 (0.00%) | 202 (61.03%) |
| Cardiovascular | 0 (0.00%) | 57 (17.22%) |

\*: Categorical variables are shown as number (percentage) and continuous variables are presented as mean ± standard deviation.

### Random Forest Analysis

Random Forest models were trained and tested to discriminate MHO from MUO, based on the predictors age, sex, body fat, visceral fat, and medication-adjusted abundances of 174 metabolites and 148 proteins. Adjustment of concentrations for common medications ensured any results being independent of potential confounding by unrelated treatments, such as hormonal contraceptives [43, 46]. Test results indicate a moderate classifier performance of AUC = 0.709, 95% CI = (0.698,0.721) and MCC = 0.394, 95% CI = (0.377,0.411). The receiver operating characteristic curve and precision-recall curve are provided in Fig. 1.

**Fig. 1.**
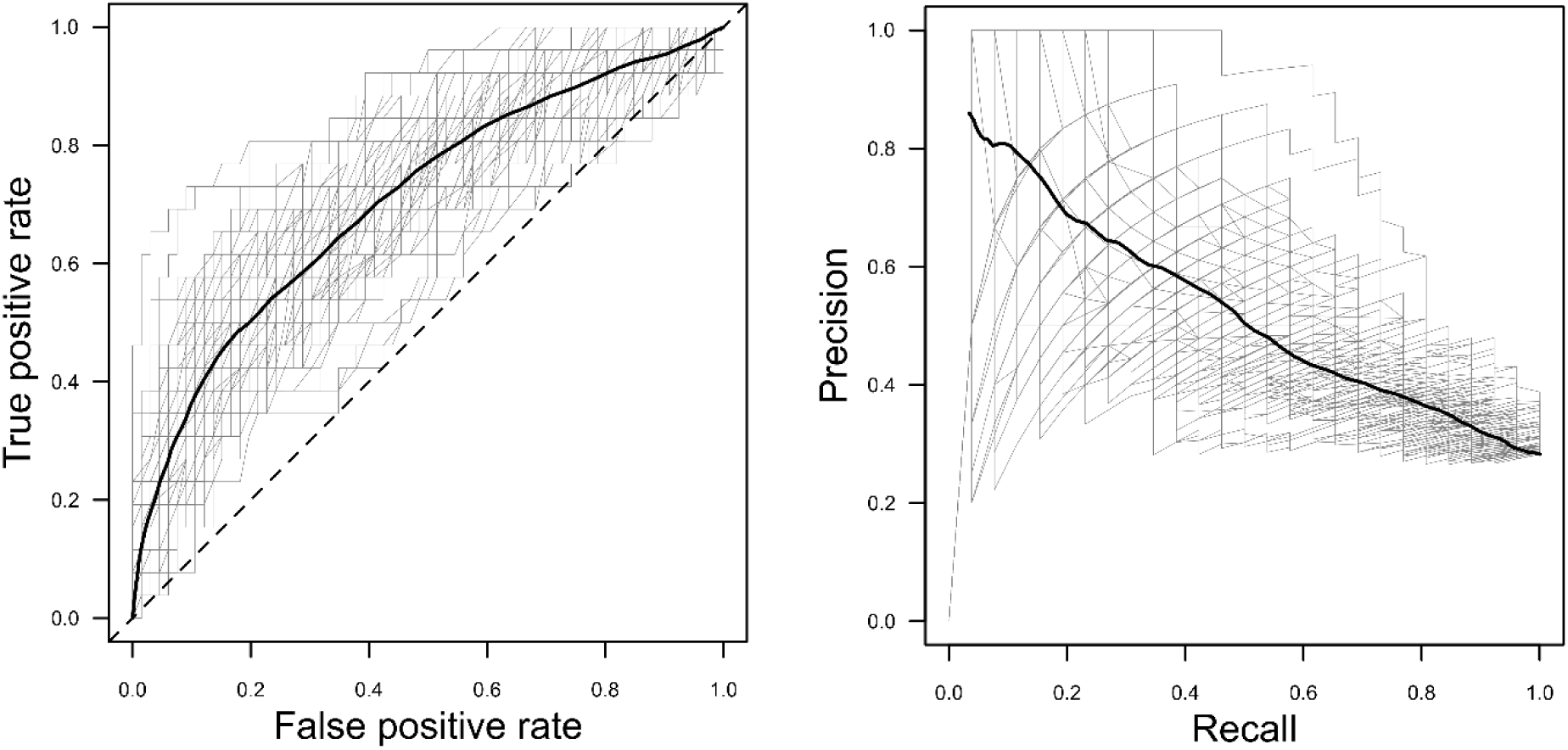
RF classification performance. ROC curve on left and precision recall curve on the right. Testing results for 100 models, each shown as gray line with average curve in black.

Individual predictors were selected based on their importance in the RF models as determined by mean decrease in Gini Index. According to selection criteria (described in the Methods section) 17 predictors were identified, including age, visceral fat, three proteins and 12 metabolites (Fig. 2a). The most relevant predictors are age, visceral fat and Apolipoprotein C-III (APOC3), followed by Insulin-like growth factor-binding protein complex acid labile subunit (IGFALS) and a phosphatidylcholine (PC aa C34:3). The distributions of the relevant predictors in both MHO and MUO phenotypes are provided in Fig. 2b. In the MHO phenotype we observe lower visceral fat and lower levels of APOC3, and higher levels of IGFALS and PC aa C34:3.

**Fig. 2.**
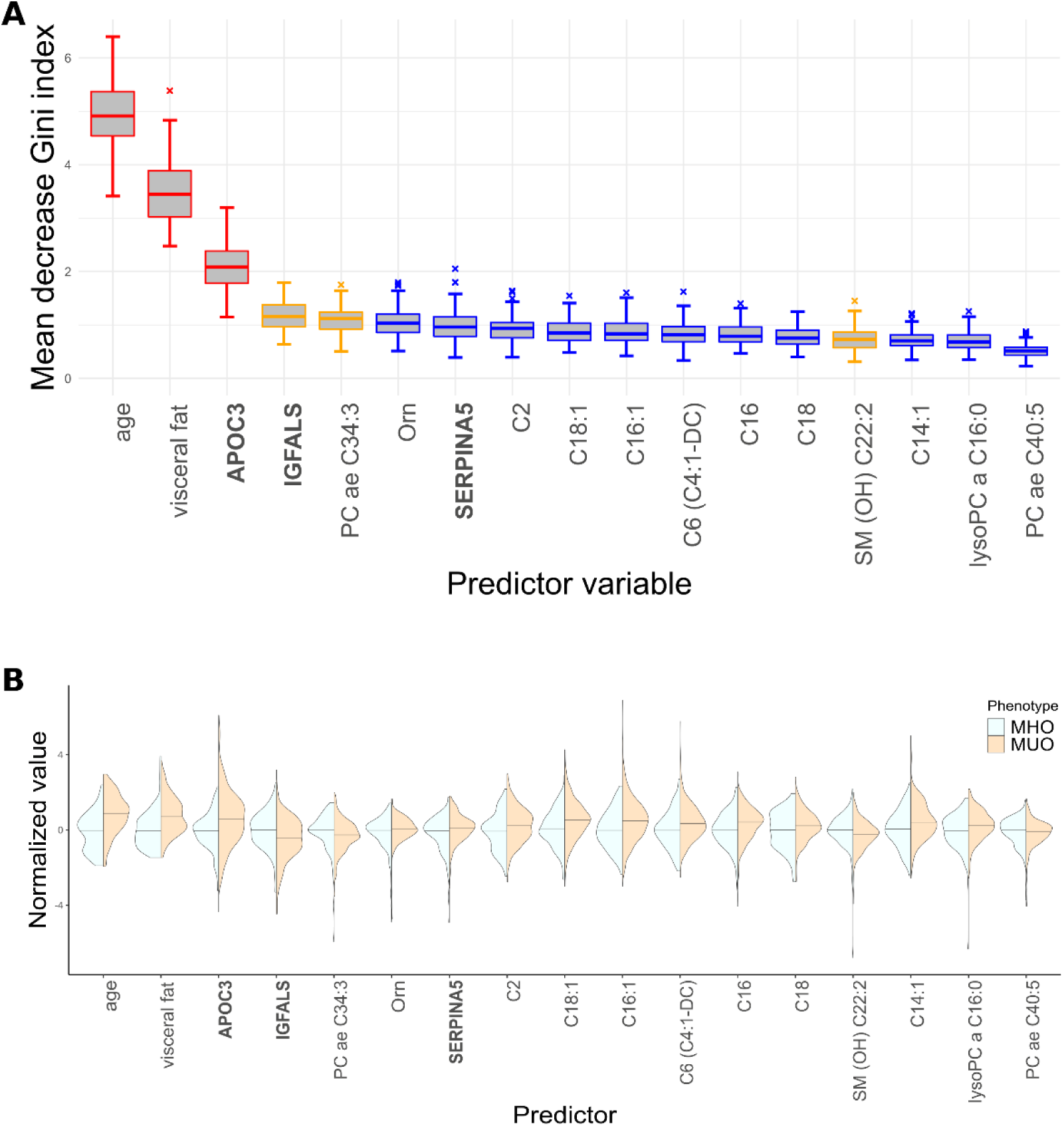
Relevant predictors for classification of MHO vs MUO phenotypes. **A** Mean decrease in Gini Index for significant predictors. Predictor significance was estimated by permutation testing, resulting in 100 p-values for each predictor variable. Box plots are color-coded by the number of times a p-value was significant for such a variable (p < 0.05): red if all 100 runs returned a significant p-value; orange when between 80 and 99 runs returned a significant p-value; blue when between 50 and 79 runs returned a significant p-value. Panels are restricted to predictors that are significant in at least 50% of all runs, and ordered by importance from left to right. **B** Violin plots presenting scaled abundance distributions for MHO and MUO groups for the significant predictors identified through RF, ordered by importance from left to right. Density plots of MHO group are scaled to have the same width, a median of zero (horizontal bar) and standard deviation of one. For comparability, the MUO group data have been scaled to the standard deviation of the MHO group. The metabolite and protein names are provided in Supplementary Tables 1 and 2. Labels for protein are in bold.

### Confirmation of relevant predictors

The relation between MHO/MUO phenotypes and the molecular predictors identified by the RF models were further investigated by separate multiple regression analyses to directly evaluate the differences in metabolite or protein concentrations between the MHO vs MUO groups independently of age, sex or body fat composition. Separate linear regression models were fitted having as response variable the medication adjusted abundances of each of the three proteins and 12 metabolites identified, and with the MHO/MUO phenotypes, sex, age and visceral fat as the explanatory variables. Detailed results are presented in Supplementary Table 4. The results confirm the association between MHO/MUO phenotypes and APOC3, plasma serine protease inhibitor (SERPINA5) and a phosphatidylcholine (PC aa C34:3), each with an adjusted p-value < 0.05. The direction of these three statistically significant associations also agrees with the observed median differences of the respective abundances in the two phenotypes (Fig. 2b). Nevertheless, in the case of SERPINA5 the association with sex is stronger than with the MHO/MUO phenotypes. The levels of insulin-like growth factor-binding protein complex acid labile subunit (IGFALS) are instead strongly associated with age and visceral fat but not with the MHO/MUO phenotypes. Further, no significant associations were observed between the remaining metabolites and MHO/MUO.

Given the strong association observed between APOC3 and MHO/MUO, we conducted a first sensitivity analysis to further investigate whether this relation could result from the intake of medication included in definition of MHO/MUO. To do so a linear regression model was fitted with APOC3 levels as response variable and MHO/MUO phenotypes and lipid modifier medication as explanatory but not with the lipid medication, indicating a robust relation between APOC3 and MHO/MUO phenotypes. A similar analysis for the PC aa C34:3 levels indicate a stronger association to statin intake than to MHO/MUO phenotypes, suggesting that the PC aa C34:3 relation to MHO/MUO phenotypes just reflects medication intake. The results are provided in Supplementary Table 5.

Given that APOC3 is involved in triglyceride transport [49], the question arises whether the relation between APOC3 levels and the MHO/MUO phenotypes just reflects serum triglyceride concentration, which is also used in the phenotype definition. To address this question, we conducted a second sensitivity analysis by fitting a logistic regression model with MHO/MUO phenotypes as response variable, and sex, age, APOC3 levels, visceral fat, and triglyceride levels as explanatory variables (Supplementary Table 6). A strong association with the phenotypes is observed for both age and triglyceride levels. A weaker but significant association is observed for APOC3 levels as well, with a change in the direction of the association after adjusting for triglyceride levels. These results indicate that APOC3 is associated with MHO/MUO phenotypes independently of triglyceride levels measured by an established clinical blood test.

## DISCUSSION

A better understanding of obesity subtypes is necessary for the development of more effective therapeutic strategies. In this study we investigated blood metabolomic and proteomic signatures to discriminate metabolically healthy from unhealthy obesity phenotypes.

Our results indicate that Apolipoprotein C-III (APOC3) is a robust molecular predictor discriminating MHO from MUO, independently of age and visceral fat, which are other strong predictors of these phenotypes. Higher abundances of this apolipoprotein are observed in the MUO phenotype. APOC3 is found in triglyceride-rich lipoproteins like chylomicrons and very-low-density lipoproteins (VLDL) and inhibits hepatic uptake of VLDL [49, 50]. This inhibition contributes to elevated levels of circulating triglycerides, influencing lipid metabolism.

Although the result is not unexpected, given that serum triglyceride levels are used in the definition of MUO, further analysis with logistic regression indicates that APOC3 levels are associated with MHO/MUO phenotypes independent of serum triglyceride levels. The findings reported here are also in agreement with previous work describing higher levels of VLDL in MUO individuals relative to MHO [51], and are consistent with previous reports indicating triglyceride levels to be a significant predictor for the transition from MHO to MUO [52]. Notably, increased APOC3 levels have been observed in women with obesity [53], and decreasing APOC3 levels have been linked to weight loss [54]. APOC3 deficiency has triglyceride lowering effects and is associated with reduced cardiovascular disease (CVD) risk [55, 56], and APOC3 polymorphisms have been associated with metabolic syndrome [57] and have an impact on the coronary artery disease risk [58]. The protein is a therapeutic target of volanesorsen, a drug approved for reduction of triglyceride levels, and is a candidate target for reducing CVD risk [49, 59].

To date, this work represents the largest study analyzing circulating proteome and metabolome signatures for the MHO/MUO phenotypes. Furthermore, the analysis includes proper adjustment of proteomics and metabolomics levels regarding medication intake.

There is no established consensus on the best criteria for MHO/MUO. Even though an established criterion was used in this work, the lack of a standard can be seen as a limitation. An alternative definition has been proposed for stratifying individuals according to mortality risk based on a large prospective study [60]. Although this new definition cannot be applied in the available CHRIS baseline study dataset, given the lack of a waist and hip circumference measurements, it would be of interest to replicate the analysis with this definition once the dataset from the ongoing CHRIS follow-up study is made available, where waist-to-hip ratios are being measured. Another limitation is the relatively limited set of blood metabolites and proteins profiled.

We foresee several opportunities for a follow-up to this work. We suggest further investigating APOC3 and VLDL levels, as well as the impact of triglyceride metabolism and transport in relation to health in obesity. The observed change in the association between APOC3 and MHO/MUO after adjustment for triglyceride levels needs further investigation. The replication of these findings in other studies and the exploration of causal relationships would be very valuable. Furthermore, it would be of great interest to investigate the relationship between changes in APOC3 and lipoprotein levels, as well as the stability of MHO phenotype and transition to MUO.

## CONCLUSION

A model based on multi-omics was developed for discriminating MHO from MUO achieving moderate performance. An analysis of the relevant features indicates APOC3 as molecular predictor for the stratification of obesity. Higher levels of plasma APOC3 are observed in MUO, independently of age, sex or medication. The results highlight the role of lipoproteins and triglyceride metabolism and/or transport in shaping metabolic health within obesity.

## DATA AVAILABILITY

Data and samples can be requested for clearly defined research via the CHRIS Portal (https://chrisportal.eurac.edu/).

## ACKNOWLEDGEMENTS

CHRIS study investigators thank all study participants, the Healthcare System of the Autonomous Province of Bolzano-South Tyrol, and all Eurac Research staff involved in the study (https://www.eurac.edu/chrisack).

## AUTHOR CONTRIBUTIONS

Conceptualization: CXW, GP, FM and FSD. Methodology: GP, CXW, JR and FSD. Software: GP and CXW. Formal analysis and investigation: GP, CXW and FSD. Resources: JR, MR, MK, FSD and PPP. Data curation: CXW, JR and MR. Writing: GP, CXW, EH, JR, MDG and FSD. Visualization: CXW. Supervision: FM and FSD. Funding acquisition: FM, MK, FSD and PPP. All authors approved the final version of the manuscript.

## ADDITIONAL INFORMATION

**Supplementary Information**

Supplementary File, PDF

