## Supplementary Information for "Characterization of metabolically healthy and unhealthy obesity through circulating proteins and metabolites"

### 1. Supplementary Table 1

In total, 188 metabolites were quantified with the Biocrates AbsoluteIDQ p180 kit, and 14 were excluded because of poor quality or high number of missing values in the CHRIS cohort.

**Supplementary Table 1.** Overview of the 174 metabolites included in the analysis.

| Analyte name | Biochemical name | Analyte class |
|---|---|---|
| ADMA | Asymmetric dimethylarginine | biogenic amines |
| Ala | Alanine | amino acids |
| alpha-AAA | alpha-Aminoadipic acid | biogenic amines |
| Arg | Arginine | amino acids |
| Asn | Asparagine | amino acids |
| Asp | Aspartate | amino acids |
| Cit | Citrulline | amino acids |
| Creatinine | Creatinine | biogenic amines |
| DOPA | Dihydroxyphenylalanine | biogenic amines |
| Gln | Glutamine | amino acids |
| Glu | Glutamate | amino acids |
| Gly | Glycine | amino acids |
| His | Histidine | amino acids |
| Histamine | Histamine | biogenic amines |
| Ile | Isoleucine | amino acids |
| Kynurenine | Kynurenine | biogenic amines |
| Leu | Leucine | amino acids |
| Lys | Lysine | amino acids |
| Met | Methionine | amino acids |
| Met-SO | Methioninesulfoxide | biogenic amines |
| Orn | Ornithine | amino acids |
| Phe | Phenylalanine | amino acids |
| Pro | Proline | amino acids |
| Putrescine | Putrescine | biogenic amines |
| SDMA | Symmetric dimethylarginine | biogenic amines |
| Ser | Serine | amino acids |
| Serotonin | Serotonin | biogenic amines |
| Spermidine | Spermidine | biogenic amines |
| Spermine | Spermine | biogenic amines |
| t4-OH-Pro | trans-4-Hydroxyproline | biogenic amines |
| Taurine | Taurine | biogenic amines |
| Thr | Threonine | amino acids |
| Trp | Tryptophan | amino acids |
| Tyr | Tyrosine | amino acids |
| Val | Valine | amino acids |
| C0 | Carnitine | acylcarnitines |
| C10 | Decanoylcarnitine | acylcarnitines |
| C10:1 | Decenoylcarnitine | acylcarnitines |
| C10:2 | Decadienylcarnitine | acylcarnitines |
| C12 | Dodecanoylcarnitine | acylcarnitines |
| C12-DC | Dodecanedioylcarnitine | acylcarnitines |
| C12:1 | Dodecenoylcarnitine | acylcarnitines |
| C14 | Tetradecanoylcarnitine | acylcarnitines |
| C14:1 | Tetradecenoylcarnitine | acylcarnitines |
| C14:1-OH | Hydroxytetradecenoylcarnitine | acylcarnitines |
| C14:2 | Tetradecadienylcarnitine | acylcarnitines |
| C14:2-OH | Hydroxytetradecadienylcarnitine | acylcarnitines |
| C16 | Hexadecanoylcarnitine | acylcarnitines |
| C16-OH | Hydroxyhexadecanoylcarnitine | acylcarnitines |

| Analyte name | Biochemical name | Analyte class |
|---|---|---|
| C16:1 | Hexadecenoylcarnitine | acylcarnitines |
| C16:1-OH | Hydroxyhexadecenoylcarnitine | acylcarnitines |
| C18 | Octadecanoylcarnitine | acylcarnitines |
| C18:1 | Octadecenoylcarnitine | acylcarnitines |
| C2 | Acetylcarnitine | acylcarnitines |
| C3 | Propionylcarnitine | acylcarnitines |
| C3-DC (C4-OH) | Hydroxybutyrylcarnitine | acylcarnitines |
| C3:1 | Propenoylcarnitine | acylcarnitines |
| C4 | Butyrylcarnitine | acylcarnitines |
| C4:1 | Butenylcarnitine | acylcarnitines |
| C5 | Valerylcarnitine | acylcarnitines |
| C5-DC (C6-OH) | Glutarylcarnitine (Hydroxyhexanoylcarnitine) | acylcarnitines |
| C5-M-DC | Methylglutarylcarnitine | acylcarnitines |
| C5-OH (C3-DC-M) | Hydroxyvalerylcarnitine (Methylmalonylcarnitine) | acylcarnitines |
| C5:1 | Tiglylcarnitine | acylcarnitines |
| C5:1-DC | Glutaconylcarnitine | acylcarnitines |
| C6 (C4:1-DC) | Hexanoylcarnitine (Fumarylcarnitine) | acylcarnitines |
| C6:1 | Hexenoylcarnitine | acylcarnitines |
| C8 | Octanoylcarnitine | acylcarnitines |
| C9 | Nonaylcarnitine | acylcarnitines |
| H1 | Hexose | sugars |
| lysoPC a C14:0 | lysoPhosphatidylcholine acyl C14:0 | glycerophospholipids |
| lysoPC a C16:0 | lysoPhosphatidylcholine acyl C16:0 | glycerophospholipids |
| lysoPC a C16:1 | lysoPhosphatidylcholine acyl C16:1 | glycerophospholipids |
| lysoPC a C17:0 | lysoPhosphatidylcholine acyl C17:0 | glycerophospholipids |
| lysoPC a C18:0 | lysoPhosphatidylcholine acyl C18:0 | glycerophospholipids |
| lysoPC a C18:1 | lysoPhosphatidylcholine acyl C18:1 | glycerophospholipids |
| lysoPC a C18:2 | lysoPhosphatidylcholine acyl C18:2 | glycerophospholipids |
| lysoPC a C20:3 | lysoPhosphatidylcholine acyl C20:3 | glycerophospholipids |
| lysoPC a C20:4 | lysoPhosphatidylcholine acyl C20:4 | glycerophospholipids |
| lysoPC a C24:0 | lysoPhosphatidylcholine acyl C24:0 | glycerophospholipids |
| lysoPC a C26:0 | lysoPhosphatidylcholine acyl C26:0 | glycerophospholipids |
| lysoPC a C26:1 | lysoPhosphatidylcholine acyl C26:1 | glycerophospholipids |
| lysoPC a C28:0 | lysoPhosphatidylcholine acyl C28:0 | glycerophospholipids |
| lysoPC a C28:1 | lysoPhosphatidylcholine acyl C28:1 | glycerophospholipids |
| PC aa C24:0 | Phosphatidylcholine diacyl C24:0 | glycerophospholipids |
| PC aa C26:0 | Phosphatidylcholine diacyl C26:0 | glycerophospholipids |
| PC aa C28:1 | Phosphatidylcholine diacyl C28:1 | glycerophospholipids |
| PC aa C30:0 | Phosphatidylcholine diacyl C30:0 | glycerophospholipids |
| PC aa C30:2 | Phosphatidylcholine diacyl C30:2 | glycerophospholipids |
| PC aa C32:0 | Phosphatidylcholine diacyl C32:0 | glycerophospholipids |
| PC aa C32:1 | Phosphatidylcholine diacyl C32:1 | glycerophospholipids |
| PC aa C32:2 | Phosphatidylcholine diacyl C32:2 | glycerophospholipids |
| PC aa C32:3 | Phosphatidylcholine diacyl C32:3 | glycerophospholipids |
| PC aa C34:1 | Phosphatidylcholine diacyl C34:1 | glycerophospholipids |
| PC aa C34:2 | Phosphatidylcholine diacyl C34:2 | glycerophospholipids |
| PC aa C34:3 | Phosphatidylcholine diacyl C34:3 | glycerophospholipids |
| PC aa C34:4 | Phosphatidylcholine diacyl C34:4 | glycerophospholipids |
| PC aa C36:0 | Phosphatidylcholine diacyl C36:0 | glycerophospholipids |
| PC aa C36:1 | Phosphatidylcholine diacyl C36:1 | glycerophospholipids |
| PC aa C36:2 | Phosphatidylcholine diacyl C36:2 | glycerophospholipids |
| PC aa C36:3 | Phosphatidylcholine diacyl C36:3 | glycerophospholipids |
| PC aa C36:4 | Phosphatidylcholine diacyl C36:4 | glycerophospholipids |
| PC aa C36:5 | Phosphatidylcholine diacyl C36:5 | glycerophospholipids |
| PC aa C36:6 | Phosphatidylcholine diacyl C36:6 | glycerophospholipids |
| PC aa C38:0 | Phosphatidylcholine diacyl C38:0 | glycerophospholipids |

| **Analyte name** | **Biochemical name** | **Analyte class** |
|---|---|---|
| PC aa C38:1 | Phosphatidylcholine diacyl C38:1 | glycerophospholipids |
| PC aa C38:3 | Phosphatidylcholine diacyl C38:3 | glycerophospholipids |
| PC aa C38:4 | Phosphatidylcholine diacyl C38:4 | glycerophospholipids |
| PC aa C38:5 | Phosphatidylcholine diacyl C38:5 | glycerophospholipids |
| PC aa C38:6 | Phosphatidylcholine diacyl C38:6 | glycerophospholipids |
| PC aa C40:1 | Phosphatidylcholine diacyl C40:1 | glycerophospholipids |
| PC aa C40:2 | Phosphatidylcholine diacyl C40:2 | glycerophospholipids |
| PC aa C40:3 | Phosphatidylcholine diacyl C40:3 | glycerophospholipids |
| PC aa C40:4 | Phosphatidylcholine diacyl C40:4 | glycerophospholipids |
| PC aa C40:5 | Phosphatidylcholine diacyl C40:5 | glycerophospholipids |
| PC aa C40:6 | Phosphatidylcholine diacyl C40:6 | glycerophospholipids |
| PC aa C42:0 | Phosphatidylcholine diacyl C42:0 | glycerophospholipids |
| PC aa C42:1 | Phosphatidylcholine diacyl C42:1 | glycerophospholipids |
| PC aa C42:2 | Phosphatidylcholine diacyl C42:2 | glycerophospholipids |
| PC aa C42:4 | Phosphatidylcholine diacyl C42:4 | glycerophospholipids |
| PC aa C42:5 | Phosphatidylcholine diacyl C42:5 | glycerophospholipids |
| PC aa C42:6 | Phosphatidylcholine diacyl C42:6 | glycerophospholipids |
| PC ae C30:0 | Phosphatidylcholine acyl-alkyl C30:0 | glycerophospholipids |
| PC ae C30:1 | Phosphatidylcholine acyl-alkyl C30:1 | glycerophospholipids |
| PC ae C30:2 | Phosphatidylcholine acyl-alkyl C30:2 | glycerophospholipids |
| PC ae C32:1 | Phosphatidylcholine acyl-alkyl C32:1 | glycerophospholipids |
| PC ae C32:2 | Phosphatidylcholine acyl-alkyl C32:2 | glycerophospholipids |
| PC ae C34:0 | Phosphatidylcholine acyl-alkyl C34:0 | glycerophospholipids |
| PC ae C34:1 | Phosphatidylcholine acyl-alkyl C34:1 | glycerophospholipids |
| PC ae C34:2 | Phosphatidylcholine acyl-alkyl C34:2 | glycerophospholipids |
| PC ae C34:3 | Phosphatidylcholine acyl-alkyl C34:3 | glycerophospholipids |
| PC ae C36:0 | Phosphatidylcholine acyl-alkyl C36:0 | glycerophospholipids |
| PC ae C36:1 | Phosphatidylcholine acyl-alkyl C36:1 | glycerophospholipids |
| PC ae C36:2 | Phosphatidylcholine acyl-alkyl C36:2 | glycerophospholipids |
| PC ae C36:3 | Phosphatidylcholine acyl-alkyl C36:3 | glycerophospholipids |
| PC ae C36:4 | Phosphatidylcholine acyl-alkyl C36:4 | glycerophospholipids |
| PC ae C36:5 | Phosphatidylcholine acyl-alkyl C36:5 | glycerophospholipids |
| PC ae C38:0 | Phosphatidylcholine acyl-alkyl C38:0 | glycerophospholipids |
| PC ae C38:1 | Phosphatidylcholine acyl-alkyl C38:1 | glycerophospholipids |
| PC ae C38:2 | Phosphatidylcholine acyl-alkyl C38:2 | glycerophospholipids |
| PC ae C38:3 | Phosphatidylcholine acyl-alkyl C38:3 | glycerophospholipids |
| PC ae C38:4 | Phosphatidylcholine acyl-alkyl C38:4 | glycerophospholipids |
| PC ae C38:5 | Phosphatidylcholine acyl-alkyl C38:5 | glycerophospholipids |
| PC ae C38:6 | Phosphatidylcholine acyl-alkyl C38:6 | glycerophospholipids |
| PC ae C40:1 | Phosphatidylcholine acyl-alkyl C40:1 | glycerophospholipids |
| PC ae C40:2 | Phosphatidylcholine acyl-alkyl C40:2 | glycerophospholipids |
| PC ae C40:3 | Phosphatidylcholine acyl-alkyl C40:3 | glycerophospholipids |
| PC ae C40:4 | Phosphatidylcholine acyl-alkyl C40:4 | glycerophospholipids |
| PC ae C40:5 | Phosphatidylcholine acyl-alkyl C40:5 | glycerophospholipids |
| PC ae C40:6 | Phosphatidylcholine acyl-alkyl C40:6 | glycerophospholipids |
| PC ae C42:0 | Phosphatidylcholine acyl-alkyl C42:0 | glycerophospholipids |
| PC ae C42:1 | Phosphatidylcholine acyl-alkyl C42:1 | glycerophospholipids |
| PC ae C42:2 | Phosphatidylcholine acyl-alkyl C42:2 | glycerophospholipids |
| PC ae C42:3 | Phosphatidylcholine acyl-alkyl C42:3 | glycerophospholipids |
| PC ae C42:4 | Phosphatidylcholine acyl-alkyl C42:4 | glycerophospholipids |
| PC ae C42:5 | Phosphatidylcholine acyl-alkyl C42:5 | glycerophospholipids |
| PC ae C44:3 | Phosphatidylcholine acyl-alkyl C44:3 | glycerophospholipids |
| PC ae C44:4 | Phosphatidylcholine acyl-alkyl C44:4 | glycerophospholipids |
| PC ae C44:5 | Phosphatidylcholine acyl-alkyl C44:5 | glycerophospholipids |
| PC ae C44:6 | Phosphatidylcholine acyl-alkyl C44:6 | glycerophospholipids |
| SM (OH) C14:1 | Hydroxysphingomyeline C14:1 | sphingolipids |

| Analyte name | Biochemical name | Analyte class |
|---|---|---|
| SM (OH) C16:1 | Hydroxysphingomyeline C16:1 | sphingolipids |
| SM (OH) C22:1 | Hydroxysphingomyeline C22:1 | sphingolipids |
| SM (OH) C22:2 | Hydroxysphingomyeline C22:2 | sphingolipids |
| SM (OH) C24:1 | Hydroxysphingomyeline C24:1 | sphingolipids |
| SM C16:0 | Sphingomyeline C16:0 | sphingolipids |
| SM C16:1 | Sphingomyeline C16:1 | sphingolipids |
| SM C18:0 | Sphingomyeline C18:0 | sphingolipids |
| SM C18:1 | Sphingomyeline C18:1 | sphingolipids |
| SM C20:2 | Sphingomyeline C20:2 | sphingolipids |
| SM C24:0 | Sphingomyeline C24:0 | sphingolipids |
| SM C24:1 | Sphingomyeline C24:1 | sphingolipids |
| SM C26:0 | Sphingomyeline C26:0 | sphingolipids |
| SM C26:1 | Sphingomyeline C26:1 | sphingolipids |

### 2. Supplementary Table 2

**Supplementary Table 2.** Overview of the 148 plasma proteins quantified using Scanning SWATH in the CHRIS cohort.

| Protein | Gene | Description |
|---|---|---|
| P02768 | *ALB* | Serum albumin |
| P02766 | *TTR* | Transthyretin |
| P19827 | *ITIH1* | Inter-alpha-trypsin inhibitor heavy chain H1 |
| P01023 | *A2M* | Alpha-2-macroglobulin |
| P01042;P01042-2 | *KNG1* | Kininogen-1 |
| P02649 | *APOE* | Apolipoprotein E |
| P01024 | *C3* | Complement C3 |
| P04196 | *HRG* | Histidine-rich glycoprotein |
| P01011 | *SERPINA3* | Alpha-1-antichymotrypsin |
| P02787 | *TF* | Serotransferrin |
| P01834 | *IGKC* | Immunoglobulin kappa constant |
| Q14624;Q14624-2 | *ITIH4* | Inter-alpha-trypsin inhibitor heavy chain H4 |
| P07360 | *C8G* | Complement component C8 gamma chain |
| P00450 | *CP* | Ceruloplasmin |
| P02647 | *APOA1* | Apolipoprotein A-I |
| P02760 | *AMBP* | Protein AMBP |
| P01008 | *SERPINC1* | Antithrombin-III |
| O95445 | *APOM* | Apolipoprotein M |
| P01031 | *C5* | Complement C5 |
| P01042-2 | *KNG1* | Isoform LMW of Kininogen-1 |
| P06396;P06396-2 | *GSN* | Gelsolin |
| P25311 | *AZGP1* | Zinc-alpha-2-glycoprotein |
| P22792 | *CPN2* | Carboxypeptidase N subunit 2 |
| Q96PD5 | *PGLYRP2* | N-acetylmuramoyl-L-alanine amidase |
| P19823 | *ITIH2* | Inter-alpha-trypsin inhibitor heavy chain H2 |
| P02675 | *FGB* | Fibrinogen beta chain |
| P02748 | *C9* | Complement component C9 |
| P05155;P05155-3 | *SERPING1* | Plasma protease C1 inhibitor |
| P43652 | *AFM* | Afamin |
| P02679;P02679-2 | *FGG* | Fibrinogen gamma chain |
| P01861 | *IGHG4* | Immunoglobulin heavy constant gamma 4 |
| P06727 | *APOA4* | Apolipoprotein A-IV |
| O14791;O14791-2;O14791-3 | *APOL1* | Apolipoprotein L1 |
| P00751 | *CFB* | Complement factor B |
| P02750 | *LRG1* | Leucine-rich alpha-2-glycoprotein |
| P20851;P20851-2 | *C4BPB* | C4b-binding protein beta chain |
| P13671 | *C6* | Complement component C6 |
| P01019 | *AGT* | Angiotensinogen |
| P35858;P35858-2 | *IGFALS* | Insulin-like growth factor-binding protein complex acid labile subunit |
| P02671 | *FGA* | Fibrinogen alpha chain |
| P04114 | *APOB* | Apolipoprotein B-100 |
| P36955 | *SERPINF1* | Pigment epithelium-derived factor |
| P07357 | *C8A* | Complement component C8 alpha chain |
| P02751 | *FN1* | Fibronectin |
| P02765 | *AHSG* | Alpha-2-HS-glycoprotein |
| P00488 | *F13A1* | Coagulation factor XIII A chain |
| A0A0C4DH29 | *IGHV1-3* | Immunoglobulin heavy variable 1-3 |
| P01602 | *IGKV1-5* | Immunoglobulin kappa variable 1-5 |
| P01619 | *IGKV3-20* | Immunoglobulin kappa variable 3-20 |

| **Protein** | **Gene** | **Description** |
|---|---|---|
| P10909;P10909-2;P10909-4;P10909-5 | *CLU* | Clusterin |
| P00746 | *CFD* | Complement factor D |
| P15814 | *IGLL1* | Immunoglobulin lambda-like polypeptide 1 |
| P00747 | *PLG* | Plasminogen |
| P08571 | *CD14* | Monocyte differentiation antigen CD14 |
| P02749 | *APOH* | Beta-2-glycoprotein 1 |
| P04217;P04217-2 | *A1BG* | Alpha-1B-glycoprotein |
| P00738 | *HP* | Haptoglobin |
| P06681 | *C2* | Complement C2 |
| P05543 | *SERPINA7* | Thyroxine-binding globulin |
| P01009 | *SERPINA1* | Alpha-1-antitrypsin |
| P04004 | *VTN* | Vitronectin |
| P05154 | *SERPINA5* | Plasma serine protease inhibitor |
| P01859 | *IGHG2* | Immunoglobulin heavy constant gamma 2 |
| P08603 | *CFH* | Complement factor H |
| P04003 | *C4BPA* | C4b-binding protein alpha chain |
| P01860 | *IGHG3* | Immunoglobulin heavy constant gamma 3 |
| P02790 | *HPX* | Hemopexin |
| P01591 | *JCHAIN* | Immunoglobulin J chain |
| P02656 | *APOC3* | Apolipoprotein C-III |
| P01877 | *IGHA2* | Immunoglobulin heavy constant alpha 2 |
| P01876 | *IGHA1* | Immunoglobulin heavy constant alpha 1 |
| P01857 | *IGHG1* | Immunoglobulin heavy constant gamma 1 |
| P01871 | *IGHM* | Immunoglobulin heavy constant mu |
| P00734 | *F2* | Prothrombin |
| A0A0J9YX35 | *IGHV3-64D* | Immunoglobulin heavy variable 3-64D |
| P02753 | *RBP4* | Retinol-binding protein 4 |
| P02746 | *C1QB* | Complement C1q subcomponent subunit B |
| P08697 | *SERPINF2* | Alpha-2-antiplasmin |
| P06310 | *IGKV2-30* | Immunoglobulin kappa variable 2-30 |
| P0C0L4 | *C4A* | Complement C4-A |
| P03952 | *KLKB1* | Plasma kallikrein |
| P07358 | *C8B* | Complement component C8 beta chain |
| P02654 | *APOC1* | Apolipoprotein C-I |
| P02774;P02774-3 | *GC* | Vitamin D-binding protein |
| P68871 | *HBB* | Hemoglobin subunit beta |
| P23142 | *FBLN1* | Fibulin-1 |
| Q9HCU4 | *CELSR2* | Cadherin EGF LAG seven-pass G-type receptor 2 |
| Q16610;Q16610-4 | *ECM1* | Extracellular matrix protein 1 |
| P02652 | *APOA2* | Apolipoprotein A-II |
| P00748 | *F12* | Coagulation factor XII |
| P27169 | *PON1* | Serum paraoxonase/arylesterase 1 |
| P80108 | *GPLD1* | Phosphatidylinositol-glycan-specific phospholipase D |
| P51884 | *LUM* | Lumican |
| P02747 | *C1QC* | Complement C1q subcomponent subunit C |
| A0A075B6I0 | *IGLV8-61* | Immunoglobulin lambda variable 8-61 |
| A0A075B6H9 | *IGLV4-69* | Immunoglobulin lambda variable 4-69 |
| P05546 | *SERPIND1* | Heparin cofactor 2 |
| P22352 | *GPX3* | Glutathione peroxidase 3 |
| P06396 | *GSN* | Gelsolin |
| P09871 | *C1S* | Complement C1s subcomponent |
| P05156 | *CFI* | Complement factor I |
| A0A0B4J1U7 | *IGHV6-1* | Immunoglobulin heavy variable 6-1 |
| A0A0J9YXX1 | *IGHV5-10-1* | Immunoglobulin heavy variable 5-10-1 |
| P01780 | *IGHV3-7* | Immunoglobulin heavy variable 3-7 |
| A0A0A0MS15 | *IGHV3-49* | Immunoglobulin heavy variable 3-49 |

| **Protein** | **Gene** | **Description** |
|---|---|---|
| P00736 | *C1R* | Complement C1r subcomponent |
| Q06033;Q06033-2 | *ITIH3* | Inter-alpha-trypsin inhibitor heavy chain H3 |
| P08519 | *LPA* | Apolipoprotein(a) |
| P08185 | *SERPINA6* | Corticosteroid-binding globulin |
| P04217 | *A1BG* | Alpha-1B-glycoprotein |
| P04278 | *SHBG* | Sex hormone-binding globulin |
| P15169 | *CPN1* | Carboxypeptidase N catalytic chain |
| P05090 | *APOD* | Apolipoprotein D |
| P10643 | *C7* | Complement component C7 |
| Q14624 | *ITIH4* | Inter-alpha-trypsin inhibitor heavy chain H4 |
| P02763 | *ORM1* | Alpha-1-acid glycoprotein 1 |
| P20742 | *PZP* | Pregnancy zone protein |
| P18428 | *LBP* | Lipopolysaccharide-binding protein |
| O43866 | *CD5L* | CD5 antigen-like |
| P02745 | *C1QA* | Complement C1q subcomponent subunit A |
| P07225 | *PROS1* | Vitamin K-dependent protein S |
| P29622 | *SERPINA4* | Kallistatin |
| O75636 | *FCN3* | Ficolin-3 |
| P01701 | *IGLV1-51* | Immunoglobulin lambda variable 1-51 |
| P01703 | *IGLV1-40* | Immunoglobulin lambda variable 1-40 |
| P06312 | *IGKV4-1* | Immunoglobulin kappa variable 4-1 |
| P01705 | *IGLV2-23* | Immunoglobulin lambda variable 2-23 |
| P49908 | *SELENOP* | Selenoprotein P |
| O75882-2 | *ATRN* | Isoform 2 of Attractin |
| Q9UGM5 | *FETUB* | Fetuin-B |
| P05452 | *CLEC3B* | Tetranectin |
| A0A0B4J1V2 | *IGHV2-26* | Immunoglobulin heavy variable 2-26 |
| P00742 | *F10* | Coagulation factor X |
| P00739 | *HPR* | Haptoglobin-related protein |
| P01599 | *IGKV1-17* | Immunoglobulin kappa variable 1-17 |
| P00740 | *F9* | Coagulation factor IX |
| P19652 | *ORM2* | Alpha-1-acid glycoprotein 2 |
| P05160 | *F13B* | Coagulation factor XIII B chain |
| A0A075B6K4 | *IGLV3-10* | Immunoglobulin lambda variable 3-10 |
| A0A0C4DH31 | *IGHV1-18* | Immunoglobulin heavy variable 1-18 |
| P80748 | *IGLV3-21* | Immunoglobulin lambda variable 3-21 |
| A0A0B4J1Y9 | *IGHV3-72* | Immunoglobulin heavy variable 3-72 |
| P23142;P23142-4 | *FBLN1* | Fibulin-1 |
| P69905 | *HBA1* | Hemoglobin subunit alpha |
| A0A075B6J9 | *IGLV2-18* | Immunoglobulin lambda variable 2-18 |
| A0A0C4DH34 | *IGHV4-28* | Immunoglobulin heavy variable 4-28 |
| B9A064 | *IGLL5* | Immunoglobulin lambda-like polypeptide 5 |
| P06276 | *BCHE* | Cholinesterase |

### 3. Supplementary Table 3

**Supplementary Table 3.** Drugs used to adjust metabolite and protein levels.

| ATC level 2 description | ATC level 4 code |
|---|---|
| DRUGS FOR ACID RELATED DISORDERS | A02BC |
| VITAMINS | A11CC |
| | A11DB |
| MINERAL SUPPLEMENTS | A12AX |
| | A12CC |
| ANTITHROMBOTIC AGENTS | B01AA |
| | B01AC |
| ANTIANEMIC PREPARATIONS | B03AA |
| | B03BB |
| CARDIAC THERAPY | C01BC |
| OTHER GYNECOLOGICALS | G02BA |
| | G02BB |
| SEX HORMONES AND MODULATORS OF THE GENITAL SYSTEM | G03AA |
| | G03AB |
| | G03AC |
| | G03CA |
| | G03FA |
| | G03HB |
| UROLOGICALS | G04BD |
| | G04CA |
| | G04CB |
| CORTICOSTEROIDS FOR SYSTEMIC USE | H02AB |
| THYROID THERAPY | H03AA |
| ANTIBACTERIALS FOR SYSTEMIC USE | J01CR |
| | J01FA |
| | J01MA |
| ANTIINFLAMMATORY AND ANTIRHEUMATIC PRODUCTS | M01AB |
| | M01AE |
| | M01AH |
| MUSCLE RELAXANTS | M03BX |
| ANTIGOUT PREPARATIONS | M04AA |
| ANALGESICS | N02AA |
| | N02BE |
| ANTIEPILEPTICS | N03AF |
| | N03AG |
| | N03AX |
| ANTI-PARKINSON DRUGS | N04BC |
| PSYCHOLEPTICS | N05AH |
| | N05BA |
| | N05CD |
| | N05CF |
| PSYCHOANALEPTICS | N06AA |
| | N06AB |
| | N06AX |
| OTHER NERVOUS SYSTEM DRUGS | N07CA |
| NASAL PREPARATIONS | R01AD |
| DRUGS FOR OBSTRUCTIVE AIRWAY DISEASES | R03AC |
| | R03AK |
| | R03BA |
| | R03BB |

| ATC level 2 description | ATC level 4 code |
|---|---|
| COUGH AND COLD PREPARATIONS | R05CB |
| ANTIHISTAMINES FOR SYSTEMIC USE | R06AE |
| | R06AX |
| OPHTHALMOLOGICALS | S01ED |
| | S01EE |

### 4. List of Linear Models

#### 4.1. Correction for Medication Usage

We correct for use of medication for the most abundant drugs taken by participants. The linar model is formulated as

`Response ~` $D_1$ `+` $D_2$ `+ ... +` $D_{55}$

where `Response` describes levels of protein or metabolite and $D_i$, $i$ = 1, 2, ..., 55, are binary variables indicating whether a participant takes drug $D_i$ or not. The 55 drugs used for adjustment are listed in Suppl. Table 3. Model residuals are taken as input for any computation in this work.

#### 4.2. Proteomics or Metabolomics Variable as Response

In this model we are interested to see if the phenotype can be used to predict levels of protein or metabolite. From the list of chosen important predictors of the random forest model, we use each of them as a response variable of a linear model, adjusting for MHO/MUO phenotypes, sex, age, and visceral fat. So the linear model equals to

```
Response ~ MHO/MUO Phenotypes + Age + Sex + Visceral fat
```

with proteins APOC3, IGFALS, SERPINA5, and metabolites PC ae C34:3, SM (OH) C22:2, PC ae C40:5, lysoPC a C16:0, Orn, C2, C18:1, C16:1, C6 (C4:1-DC), C16, C18, C14:1 implemented as response variables.

**Supplementary Table 4.** Linear model features.

| Response | MHO/MUO Phenotypes | | | Age | | | Sex:Female | | | Visceral fat | | |
|---|---|---|---|---|---|---|---|---|---|---|---|---|
| | **β** | **P** | **$P_{adj}$** | **β** | **P** | **$P_{adj}$** | **β** | **P** | **$P_{adj}$** | **β** | **P** | **$P_{adj}$** |
| APOC3 | 0.2590 | 6.15E-07 | **1.85E-06** | -0.0004 | 8.08E-01 | 1.00 | -0.1201 | 4.59E-02 | 1.38E-01 | -0.0074 | 3.41E-01 | 1.00 |
| IGFALS | -0.0500 | 2.82E-01 | 8.47E-01 | -0.0120 | 3.62E-13 | **1.09E-12** | -0.0335 | 5.38E-01 | 1.00 | -0.0190 | 7.43E-03 | **2.23E-02** |
| SERPINA5 | 0.2901 | 1.28E-03 | **3.83E-03** | -0.0002 | 9.53E-01 | 1.00 | -0.3598 | 6.54E-04 | **1.96E-03** | -0.0028 | 8.35E-01 | 1.00 |
| PC ae C34:3 | -0.1825 | 2.52E-03 | **3.03E-02** | 0.0037 | 7.42E-02 | 8.90E-01 | -0.0456 | 5.18E-01 | 1.00 | -0.0112 | 2.20E-01 | 1.00 |
| SM (OH) C22:2 | -0.1279 | 8.64E-03 | 1.04E-01 | 0.0049 | 3.72E-03 | **4.46E-02** | 0.2843 | 8.07E-07 | **9.69E-06** | -0.0011 | 8.86E-01 | 1.00 |
| PC ae C40:5 | -0.0655 | 1.67E-01 | 1.00 | 0.0045 | 5.75E-03 | 6.90E-02 | 0.0121 | 8.27E-01 | 1.00 | -0.0003 | 9.67E-01 | 1.00 |
| lysoPC a C16:0 | 0.0900 | 1.39E-02 | 1.67E-01 | 0.0044 | 4.48E-04 | **5.38E-03** | -0.2205 | 3.67E-07 | **4.40E-06** | -0.0106 | 5.67E-02 | 6.80E-01 |
| Orn | 0.0788 | 1.39E-01 | 1.00 | 0.0067 | 3.11E-04 | **3.74E-03** | -0.1302 | 3.70E-02 | 4.44E-01 | 0.0027 | 7.42E-01 | 1.00 |
| C2 | 0.0670 | 1.76E-01 | 1.00 | 0.0106 | 1.10E-09 | **1.32E-08** | -0.0218 | 7.08E-01 | 1.00 | 0.0033 | 6.65E-01 | 1.00 |
| C18:1 | 0.0491 | 1.86E-01 | 1.00 | 0.0087 | 2.88E-11 | **3.46E-10** | -0.0813 | 6.24E-02 | 7.49E-01 | 0.0076 | 1.77E-01 | 1.00 |
| C16:1 | 0.0641 | 6.29E-02 | 7.54E-01 | 0.0072 | 2.69E-09 | **3.22E-08** | 0.0983 | 1.52E-02 | 1.82E-01 | 0.0121 | 2.15E-02 | 2.58E-01 |
| C6 (C4:1-DC) | 0.1340 | 5.81E-03 | 6.97E-02 | 0.0074 | 1.08E-05 | **1.29E-04** | 0.1322 | 2.01E-02 | 2.41E-01 | 0.0128 | 8.16E-02 | 9.80E-01 |
| C16 | 0.0737 | 6.38E-02 | 7.66E-01 | 0.0077 | 3.70E-08 | **4.44E-07** | -0.1170 | 1.22E-02 | 1.47E-01 | 0.0053 | 3.80E-01 | 1.00 |
| C18 | -0.0253 | 5.21E-01 | 1.00 | 0.0103 | 2.35E-13 | **2.83E-12** | -0.1952 | 2.82E-05 | 3.38E-04 | 0.0009 | 8.83E-01 | 1.00 |
| C14:1 | 0.1184 | 4.98E-02 | 5.97E-01 | 0.0112 | 1.07E-07 | **1.29E-06** | -0.0115 | 8.70E-01 | 1.00 | 0.0071 | 4.41E-01 | 1.00 |

β, regression coefficient; P, p-value; $P_{adj}$, p-value adjusted for multiple hypothesis testing (Bonferroni for n=3 proteins and n=12 metabolites, respectively). Significant adjusted p-values ($P_{adj}$<0.05) are highlighted in bold.

#### 4.3. Linear Model of APOC3 and PC ae C34:3 Including Correction for Medication

Based on the results shown in Suppl. Table 4 we have chosen APOC3 and PC ae C34:3 for further investigation if these response variables are dependent on medications related to the phenotype definitions. For APOC3, given the relation to lipid transport/metabolism, we investigated association for any lipid modifying agent. For PC ae C34:3 we selected two specific ATC codes (C10AA and A10BA) based on previous analysis on the association between metabolites and medication in the CHRIS metabolomics collection (unpublished results).

We therefore investigated the following two models:

```
APOC3        ~ MHO/MUO Phenotypes + Age + Sex + Visceral Fat + D_C10
PC ae C34:3 ~ MHO/MUO Phenotypes + Age + Sex + Visceral Fat + D_C10AA + D_A10BA
```

Here, $D_{C10}$ indicates use of medication with ATC code C10 (lipid modifying agents); $D_{C10AA}$ indicates use of medication with ATC code C10AA (HMG CoA reductase inhibitors); $D_{A10BA}$ indicates use of medication with ATC code A10BA (biguanides). No correction for multiple hypothesis testing was applied to these two models.

**Supplementary Table 5.** Linear model features.

| | MHO/MUO Phenotypes | | Age | | Sex:Female | | Visceral fat | | Drug 1 | | | Drug 2 | | |
|---|---|---|---|---|---|---|---|---|---|---|---|---|---|---|
| **Response** | **β** | **P** | **β** | **P** | **β** | **P** | **β** | **P** | | **β** | **P** | | **β** | **P** |
| APOC3 | 0.2526 | **1.36E-06** | -0.0008 | 6.46E-01 | -0.1219 | **4.28E-02** | -0.0075 | 3.37E-01 | $D_{C10}$ | 0.0723 | 3.00E-01 | - | - | - |
| PC ae C40:5 | -0.0596 | 2.10E-01 | 0.0047 | **3.91E-03** | 0.0160 | 7.73E-01 | -0.0003 | 9.62E-01 | $D_{C10AA}$ | -0.2369 | **6.09E-03** | $D_{A10BA}$ | -0.2646 | 5.27E-02 |

β, regression coefficient; P, p-value (significant p-values ($P<0.05$) are highlighted in bold).

#### 4.4. Logistic Regression Model to Predict the Phenotype using APOC3 and Triglyceride Levels

In order to explore the dependency on triglyceride and APOC3 levels on the MHO/MUO phenotypes, we chose to investigate two logistic regression models. No correction for multiple hypothesis testing was applied to these two models.

```
MHO/MUO Phenotypes ~ Age + Sex + Visceral Fat + APOC3
MHO/MUO Phenotypes ~ Age + Sex + Visceral Fat + APOC3 + Triglyceride
```

**Supplementary Table 6.** Logistic regression model features.

| | **Age** | | **Female** | | **Visceral fat** | | **APOC3** | | **Triglyceride** | |
|---|---|---|---|---|---|---|---|---|---|---|
| **Response** | **β** | **P** | **β** | **P** | **β** | **P** | **β** | **P** | **β** | **P** |
| MHO/MUO Phenotypes | 0.0464 | **3.88E-06** | 0.0144 | 9.66E-01 | 0.1202 | **1.07E-02** | 1.2574 | **1.43E-05** | - | - |
| MHO/MUO Phenotypes | 0.0637 | **2.34E-08** | 0.1197 | 7.41E-01 | 0.0703 | 1.44E-01 | -1.1684 | **7.70E-03** | 0.0358 | **5.73E-13** |

β, regression coefficient; P, p-value (significant p-values ($P<0.05$) are highlighted in bold).
